# Prevalence, Characteristics, and Associated Factors of Workplace Violence Against Healthcare Professionals in India: A Systematic Review and Meta-analysis

**DOI:** 10.1101/2020.01.01.20016295

**Authors:** Md Mahbub Hossain, Rachit Sharma, Samia Tasnim, Gulam Muhammed Al Kibria, Abida Sultana, Tanjul Saxena

**Author notes:** **Authors’ contributions:** MMH conceptualized the review. MMH, RS, ST, and GMAK conducted the literature review and data extraction. MMH conducted the statistical analysis. MMH and AS conducted the narrative synthesis and interpretation. MMH drafted the first version of the manuscript, which was critically revised by TS and edited by MMH. TS supervised the methodological aspects. All authors reviewed, edited, and approved the submitted version of the manuscript. **Conflict of interest:** None. **Funding:** No funding was received at any stage of preparing this manuscript. **Ethical issues:**No ethical approval was required as it is a secondary synthesis of published studies.

## Abstract

**Background:** Workplace violence (WPV) against doctors, nurses, and other healthcare professionals is a growing public health concern affecting health systems in low- and middle-income countries. In India, incidents of WPV against health workforce have become common in recent years. However, there is no synthesized evidence on the nationwide burden of WPV in healthcare. This study aims to systematically evaluate the current evidence on the prevalence, characteristics, and associated factors of WPV against healthcare professionals in India.

**Methods:** We systematically searched seven major databases and additional sources to retrieved published literature on WPV in India. We included empirical studies without any time restriction, which reported the prevalence of WPV against healthcare professionals in India. Following data extraction, we estimated the pooled prevalence of WPV using random-effects model in meta-analysis. The characteristics and associated factors of WPV were narratively synthesized from these empirical studies.

**Results:** Among 1065 articles retrieved from multiple sources, 15 studies met our inclusion and exclusion criteria. The pooled prevalence of WPV among 2849 participants in those 15 studies was 63% (95% confidence interval [CI], 54%-72%). In the subgroup analyses, the pooled prevalence for male and female was 64% (95% CI, 51%-78%) and 54% (95% CI, 47%-62%) respectively. Moreover, pooled prevalence in 12-months and lifetime was 55% (95% CI, 43%-67%) and 69% (95% CI, 61%-78%) respectively. Among the participants, the prevalence of verbal violence (52%; 95% CI, 45%-60%) was higher than physical violence (8%; 95% CI, 5%-11%). The emergency department was a common location of WPV, whereas the patients’ attendants were perpetrators in most studies. Major factors associated with WPV included ineffective patient-provider communication, less experience of the healthcare professionals, overcrowding, shortage of resources, long waiting hours, lack of security measures, dissatisfaction about health services, high cost of care, the negative role of media, and other socio-behavioral problems among the patients and healthcare professionals.

**Conclusion:** WPV is highly prevalent among healthcare professionals in India. Critical challenges within the healthcare and social context necessitate further research, better policymaking, and multipronged interventions to address the same and prevent WPV against healthcare professionals in India.

## 1 Background

Workplace violence (WPV) against healthcare professionals is a widespread and persistent public health concern around the world.^1^ National Institute for Occupational Safety and Health (NIOSH) at Centers for Disease Control and Prevention (CDC) defined WPV as “violent acts (including physical assaults and threats of assaults) directed toward persons at work or on duty.”^2^ WPV includes but not limits into abusive behavior toward authority, verbal abuse, threatening, physical assault, sexual harassment, and racial harassment.^3^ According to the Occupational Safety and Health Administration (OSHA), nearly 75% of 25,000 workplace assaults were reported in the health care and social service settings every year.^4^ They have also found that workers in health care settings are four times more likely to be harassed at work than the workers in private industry.^4^ WPV is reported as one of the major causes of death at the workplace among healthcare professionals.^5^ Worldwide, 8% to 38% of health workers experience some form of violence at some point in their careers.^1^ In addition, the prevalence of physical aggression ranges from 35-71% and of non-physical violence ranges from 38-90%.^6^ These statistics may not reflect the real magnitude of the dismal situation because WPV workplace is often under-reported. A wide range of socio-cultural factors may contribute to such under-reporting, which include perceiving WPV as a commonly occurring incident among healthcare professionals, feeling discouraged due to complex reporting systems, or a lack of interest of the hospital administration who primarily emphasizes on organizational outcomes rather than the safety and wellbeing of the health workforce.^7,8^ Despite these issues related to under-reporting, several studies have reported how WPV has impacted the physical and mental health of the healthcare professionals,^9^ their attitude to the profession and related responsibilities,^10^ communication with patients and peers in the workplace,^11^ delivery of health services in high-risk environment,^12^ quality of care,^13,14^ and health systems performances.^4,15^

Although WPV is a global health problem, recent studies have reported a higher magnitude of WPV in low and middle-income countries (LMICs).^3,16–18^ Critical factors including severe scarcity of human resources for health,^19^ overworking and burnout in overcrowded hospitals,^20^ lack of adequate facilities and logistics to deliver health services,^19,21^ pre-existing socio-behavioral problems of the patients and healthcare providers,^16,19,22^ high cost of availing health services,^23,24^ lack of appropriate security measures,^25,26^ and many more, are found to be associated with WPV in resource-constrained contexts. A high burden WPV is reported in China where about one-third of the doctors have faced some violence and thousands have been reported to be assaulted viciously.^24,27^ Similar incidents of violent outrage towards doctors have been reported from primary to tertiary care services in Bangladesh,^28^ Nepal,^29^ Pakistan,^25,30^ and Myanmar.^31^ As a consequence of these violent outbursts, many health care professionals were brutally injured, and few even lost their lives.^32^

India has a high burden of WPV similar to other LMICs. The Indian Medical Association (IMA) has reported that 75% of doctors face either physical or verbal abuse at some point of their career.^33,34^ Being one of the most densely populated countries with a severe paucity of resources to meet the ever-increasing burden of diseases at the population-level, the incidents of WPV is increasing in India over the past few years.^33,35^ In 2017, after four separate incidents of violence against junior doctors in government hospitals in Mumbai, India, more than 2000 doctors from 17 government hospitals went on strike demanding safety at workplace.^36^ In 2019, more than a thousand doctors in West Bengal, India have undergone a 7-day strike after several violent attacks on doctors at their workplaces.^37^ This regional strike received broader public attention when doctors all over India held a nationwide strike ceasing all non-emergency medical services demanding workplace safety.^38^ Such outburst of the medical community highlights the severity of this problem in the context of India.

Despite these ongoing discourses on WPV against healthcare professionals in India, there is a lack of consolidated evidence to examine the actual magnitude of this critical problem within the health system of India. Earlier studies have reported incidents of WPV in different hospitals,^39–41^ but there is no systematic review reporting the nationwide prevalence and characteristics of WPV in India. In addition, it is essential to evaluate the factors associated with WPV to develop preventive and protective measures addressing this complex public health problem. The objective of this article is to systematically review and meta-analyze the prevalence, characteristics, and factors associated with WPV against healthcare professionals in India.

## 2 Methods

### 2.1 Conducting The Review

In this systematic review, we aimed to evaluate the empirical studies and estimate the pooled prevalence of WPV among healthcare professionals in India. Further, we evaluated the prevalence of two major types of WPV, which is verbal and physical violence separately. Earlier studies conducted in similar socio-economic contexts have shown varying characteristics of WPV provide better insights on the magnitude and impacts of the problem.^3,16,19,25,28,29^ These characteristics include the location, time, frequency, and magnitude of WPV; impacts of WPV on the victims; perpetrators of WPV and measures following incidents of WPV. We examined the characteristics of WPV within a broader scope to better understand the scenario in the healthcare and societal context of India. Lastly, we evaluated factors associated with WPV as reported by the participants in these empirical studies to synthesize the evidence on the aggravating factors of WPV against healthcare professionals in India. The protocol for this review is registered in the International Prospective Register for Systematic Reviews-PROSPERO (CRD42019147723).

### 2.2 Search Strategy

This systematic review was conducted following the Preferred Reporting Items for Systematic Reviews and Meta-Analysis (PRISMA) guidelines.^42^ Studies indexed in MEDLINE, EMBASE, CINAHL, PsycINFO, Health Policy Reference Center, ERIC, and Scopus databases were searched using specific keywords as shown in Table 1. In this searching process, the keywords were used both as subject headings and general keywords with appropriate Boolean operators. No time restriction was imposed in the literature searching process. Three authors developed the search strategy and working protocol for this review prior to conducting this review. All the databases were searched on August 10, 2019, for the last time.

**Table 1:**
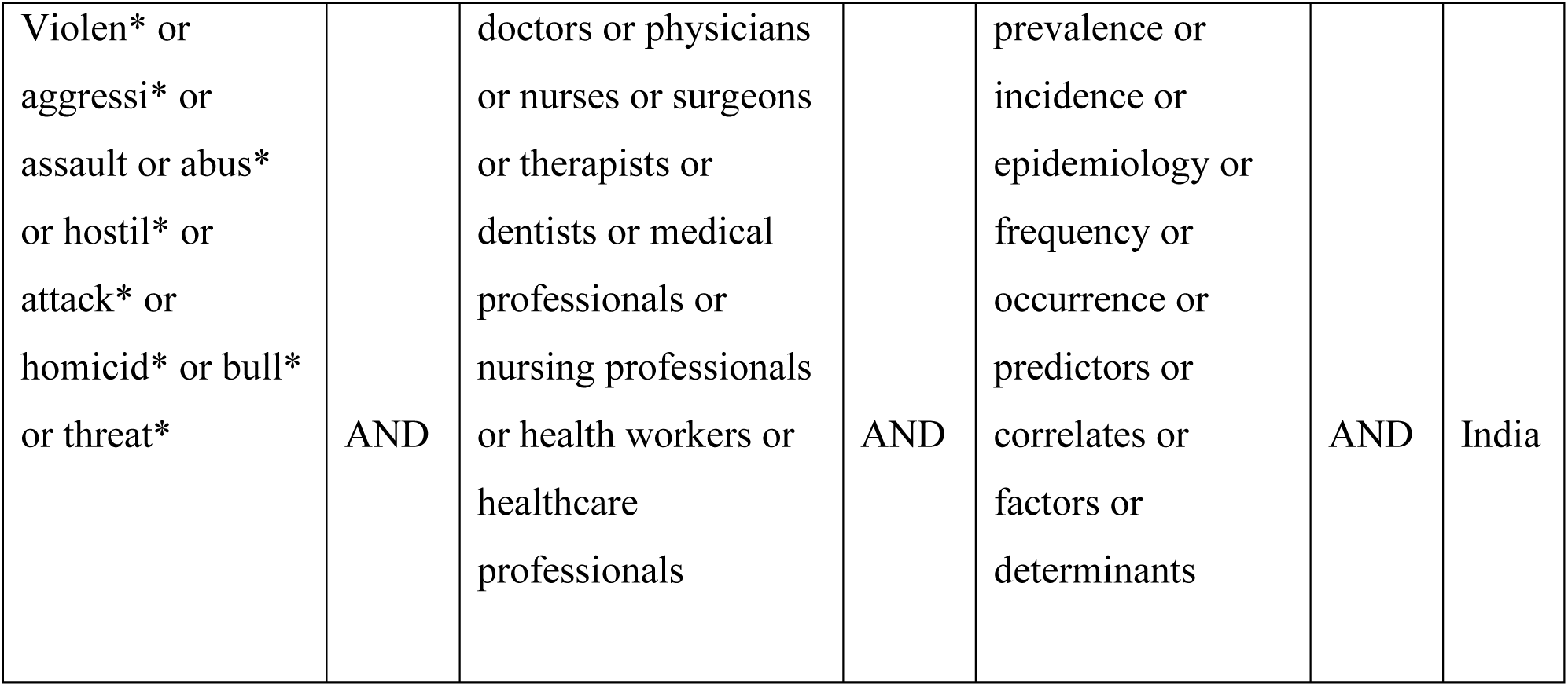
Keywords For Database Searching.

Further, we communicated with experts, healthcare researchers, and practitioners working in India to retrieve any studies as per their knowledge in the field. We also examined the citing articles of the earlier studies on this topic and the references of studies included in this review. Furthermore, we communicated with the authors of included studies to clarify doubts and retrieve missing information in the published articles. In this way, we attempted to retrieve potential studies from all these additional sources to ensure the recruited literature is inclusive and extensive enough to synthesize adequate evidence on WPV against health workforce in India.

### 2.3 Literature Screening, Inclusion and Exclusion Criteria

Citations retrieved through searching the databases and additional sources were uploaded to RefWorks^43^ and Rayyan QCRI^44^ tools for reference management and screening the citations respectively. Two authors independently conducted the screening process based on the pre-specified inclusion and exclusion criteria.

We included articles in this review if they were:

a. empirical in nature,
b. published in a peer-reviewed journal,
c. conducted among any of the health professional groups working in India,
d. conducted in quantitative or mixed methods reporting at least the prevalence of WPV in the given population as assessing the same was the primary objective of this review,
e. studies published in the English language only.

The articles were excluded from this systematic review if they were:

a. not empirical studies in nature (e.g., editorials, letters, opinions, and reviews),
b. conducted outside India,
c. did not include healthcare professionals as study participants
d. were not published in a peer-reviewed journal,
e. did not report the prevalence of WPV among the health professionals,
f. published in languages other than English.

At the end of blinded primary screening by two authors, conflicts on recruitment of citations were discussed in the presence of a third author to reach a consensus.

### 2.4 Data Extraction and Synthesis

Two authors independently re-evaluated the full texts of the finally recruited articles and extracted data in a pre-designed codebook for data extraction. This codebook had following domains to extract data: a) location and time of the study, b) types of instruments used to assess WPV, c) recruitment sites and strategies, d) prevalence of WPV in the study, e) any characteristics of WPV reported by the participants, f) associated factors of WPV.

The extracted data were further reviewed by two more authors to identify and resolve inconsistencies. Further, we included the key findings extracted from individual studies and organized the same in a tabulated format. Quantitative data on the prevalence of WPV were considered for meta-analysis, and remaining data on characteristics and associated factors of WPV were used for narrative synthesis.

### 2.5 Statistical Analysis

To estimate the pooled prevalence of WPV in the meta-analysis, we used the Der-Simonian and Laird’s random model with 95% confidence interval (CI).^45^ Stata Version 15 software (StataCorp, College Station, TX) was used for statistical analyses.^46^ Prevalence rates were obtained from all the recruited studies with priority to the overall prevalence of WPV followed by itemized prevalence for different types of WPV among the study participants. In cases of absent values for overall prevalence, the highest reported rate of WPV in the sub-groups was considered in the analysis. Further, positive cases were divided by the total number of sample in intercept-only random effects logistic regression model fitted to obtain the pooled estimate of prevalence. To estimate the prevalence in the context of different variables, we conducted subgroup analysis for gender, sample size below and above 200, publication before and after 2016, prevalence in last 12 months and without any time restriction or lifetime, and major types of WPV. To estimate the pooled prevalence across studies, “metaprop_one” program in Stata 15.0 (College Station, TX) was used, which provides better estimates without excluding 0% or 100% proportions.^47^ Moreover, this process reports study-specific and pooled confidence intervals within admissible values, I2 statistics which facilitates interpreting the heterogeneity of the studies, and P-value from Q-statistics to evaluate the between-study variability.

### 2.6 Quality Assessment of the Included Studies

In this systematic review, we used the National Heart, Lung, and Blood Institute (NHLBI) Study Quality Assessment Tool for observational cohort and cross-sectional studies to assess the methodological quality of the included studies.^48^ The NHLBI tool has 14 items which are used in previous systematic reviews of observational studies.^49,50^ Each item is assessed through a question where the possible answers are yes, no, or other (cannot determine, not applicable, or not reported). The NHLBI tool enables to score and weigh the evidence to rate the overall quality of each study as ‘good’, ‘fair’, or ‘poor’ indicating the validity of the findings in the respective study.

## 3 Results

A total of 1026 articles were found through searching seven databases. Moreover, we found 39 additional articles from additional sources (Figure 1). Therefore, the total number of articles primarily considered in this review was 1065. Further, we removed 417 duplicate articles and evaluated the titles and abstracts of the remaining 648 articles based on the inclusion or exclusion criteria, as described earlier. At the end of this scrutiny, 614 articles that did not meet our selection criteria were filtered out. Further, we evaluated the full texts of 34 remaining articles, among which 19 were excluded due to mismatched study objectives and characteristics conflicting with this review. Finally, we found 15 full-text articles which fulfilled all the criteria for this systematic review (Table 2).

**Table 2:**
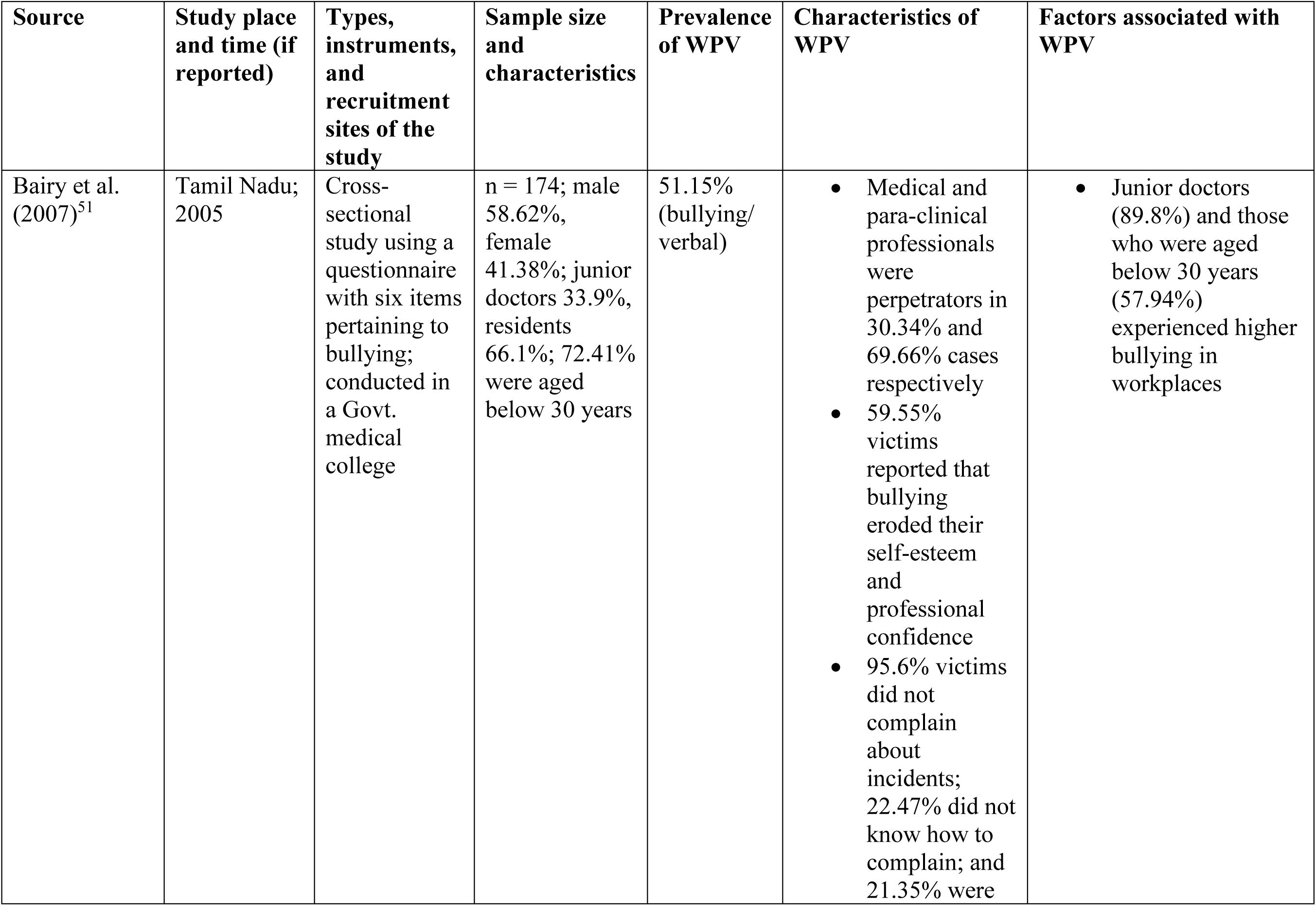
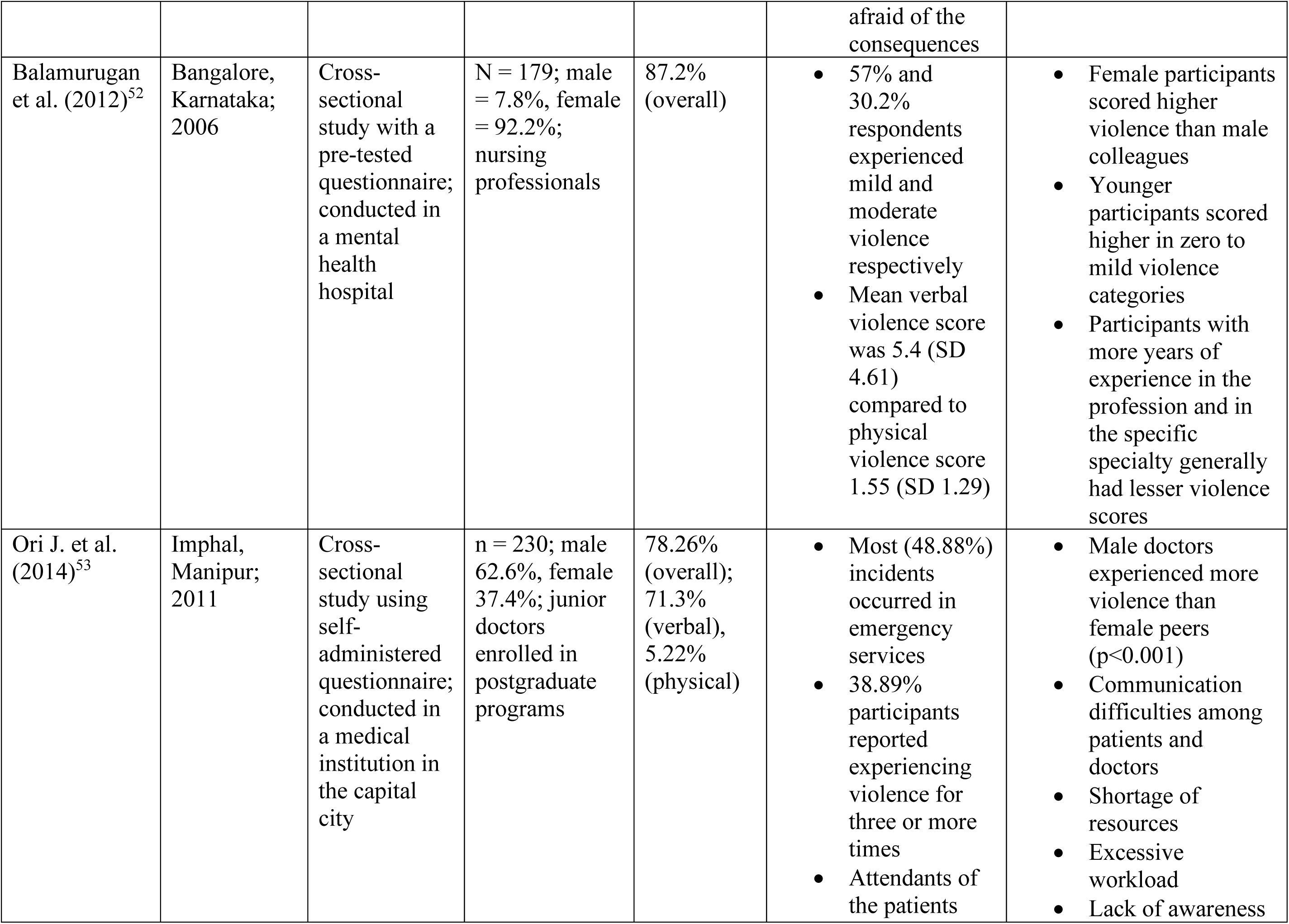
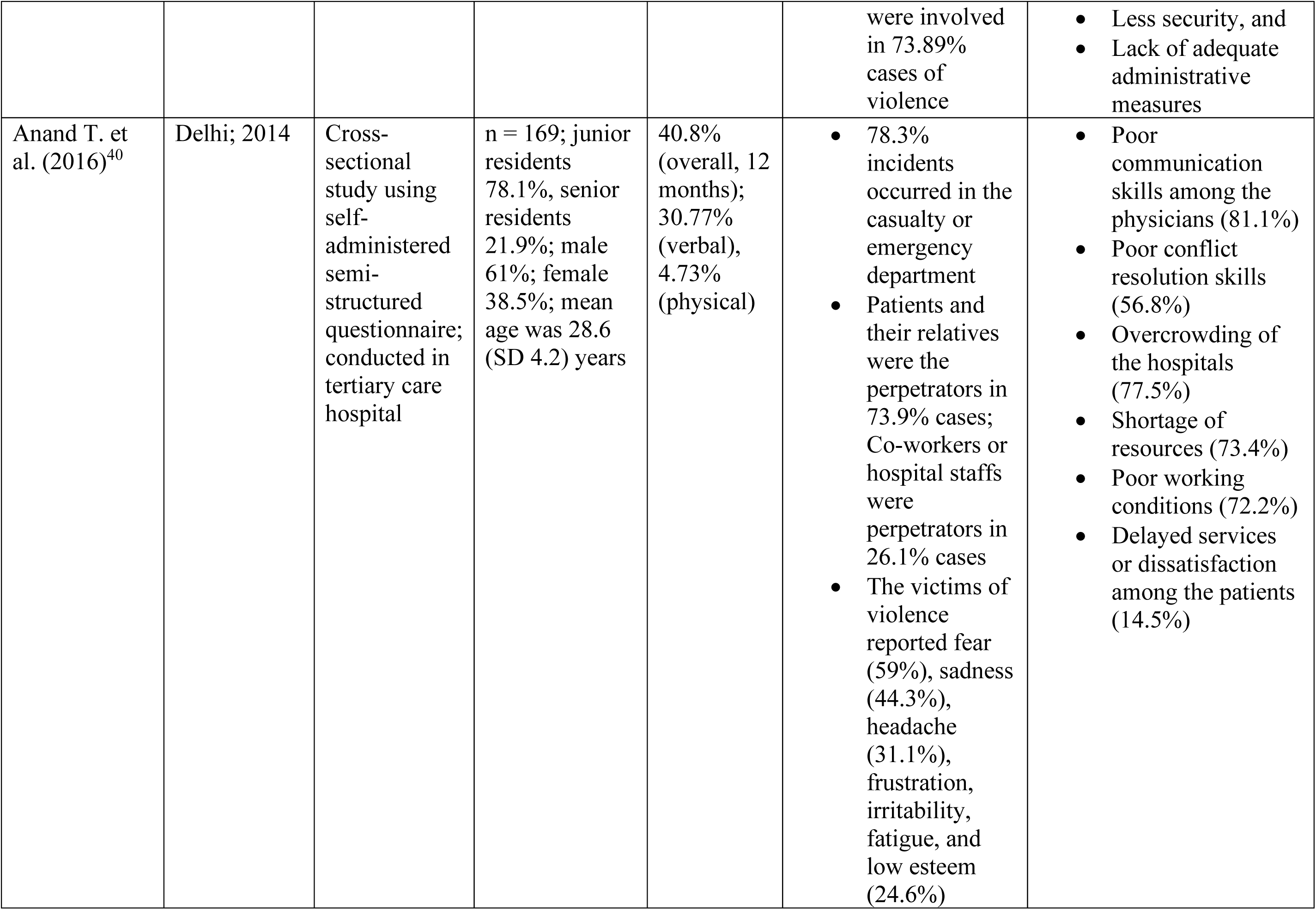
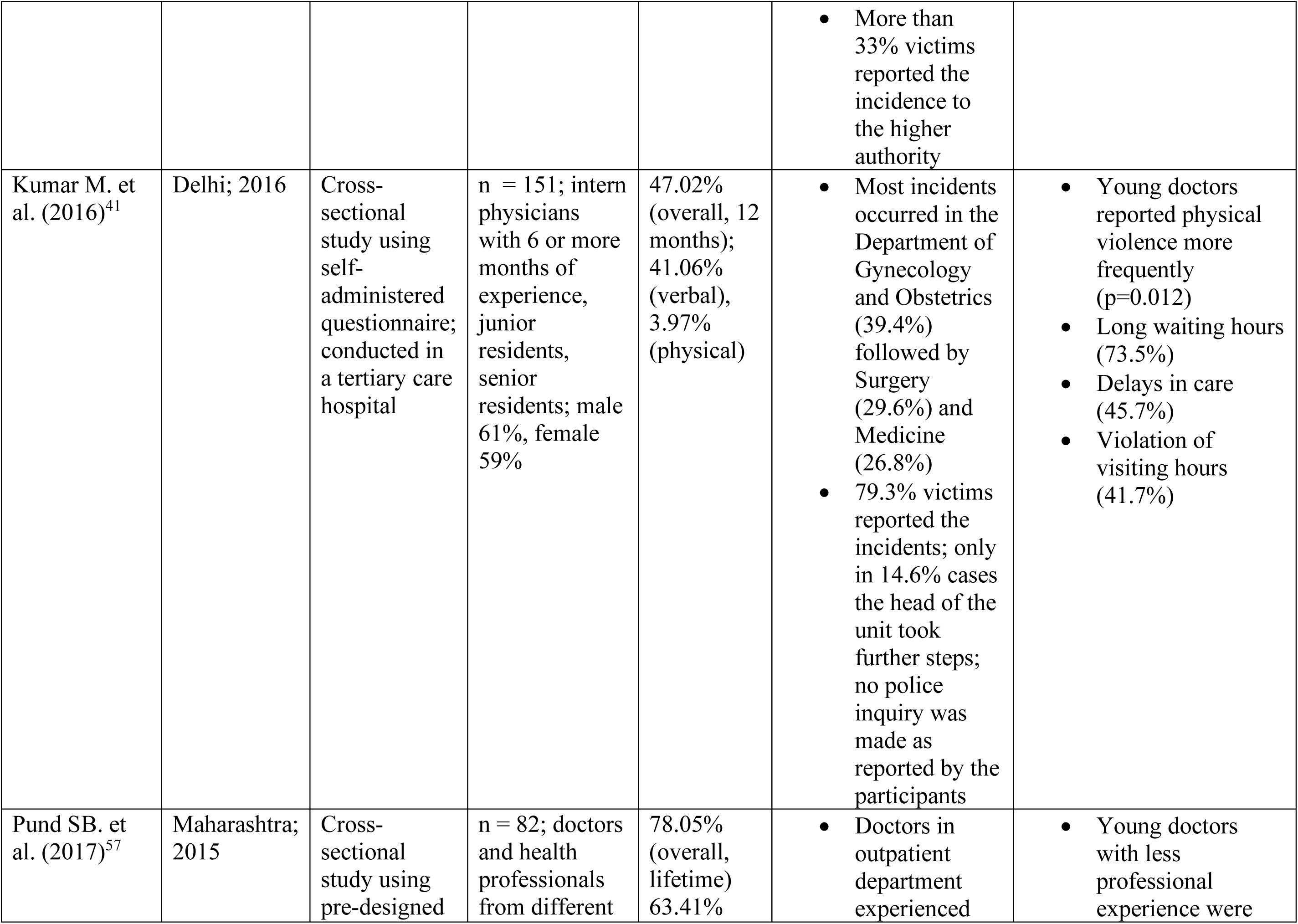
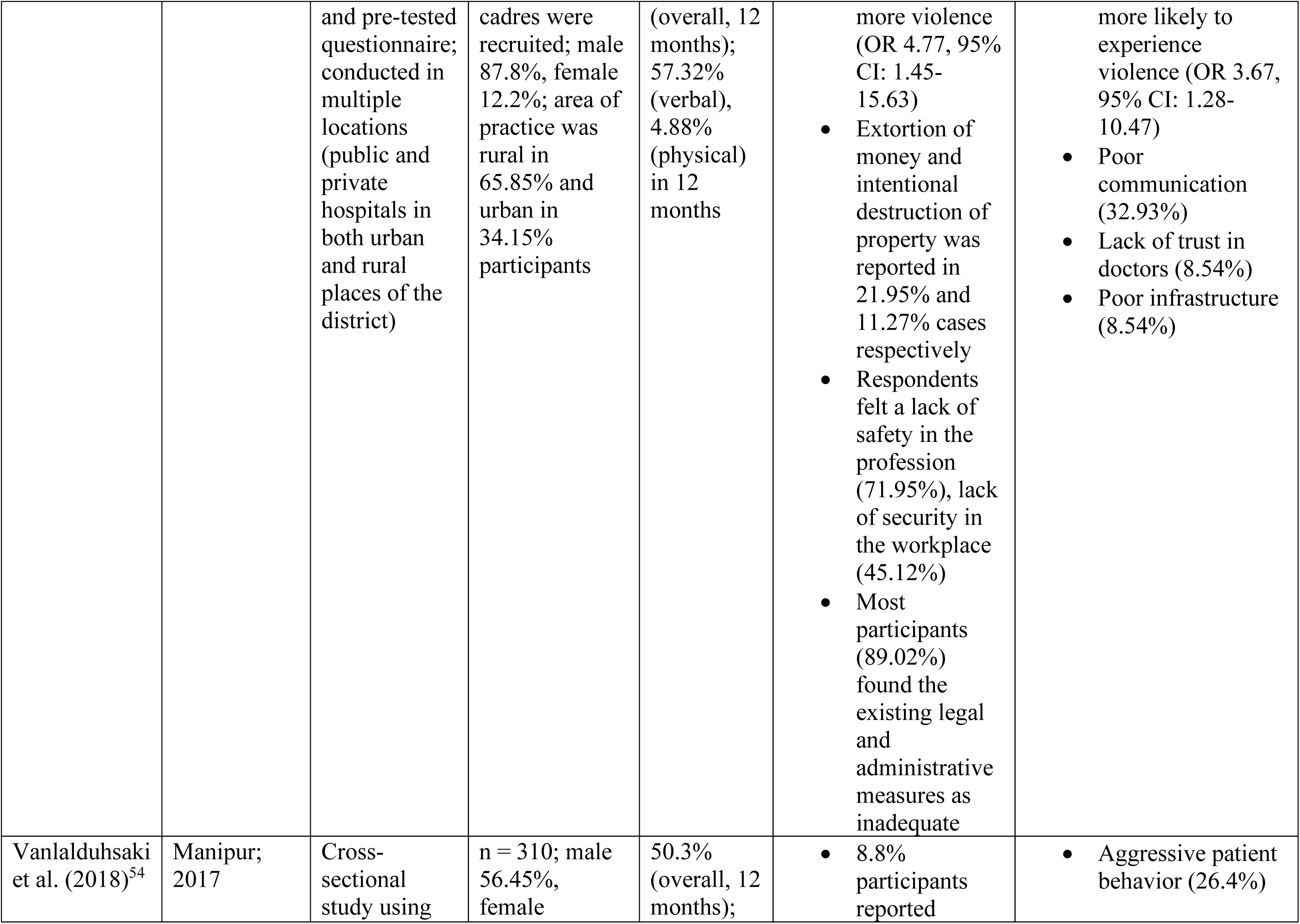
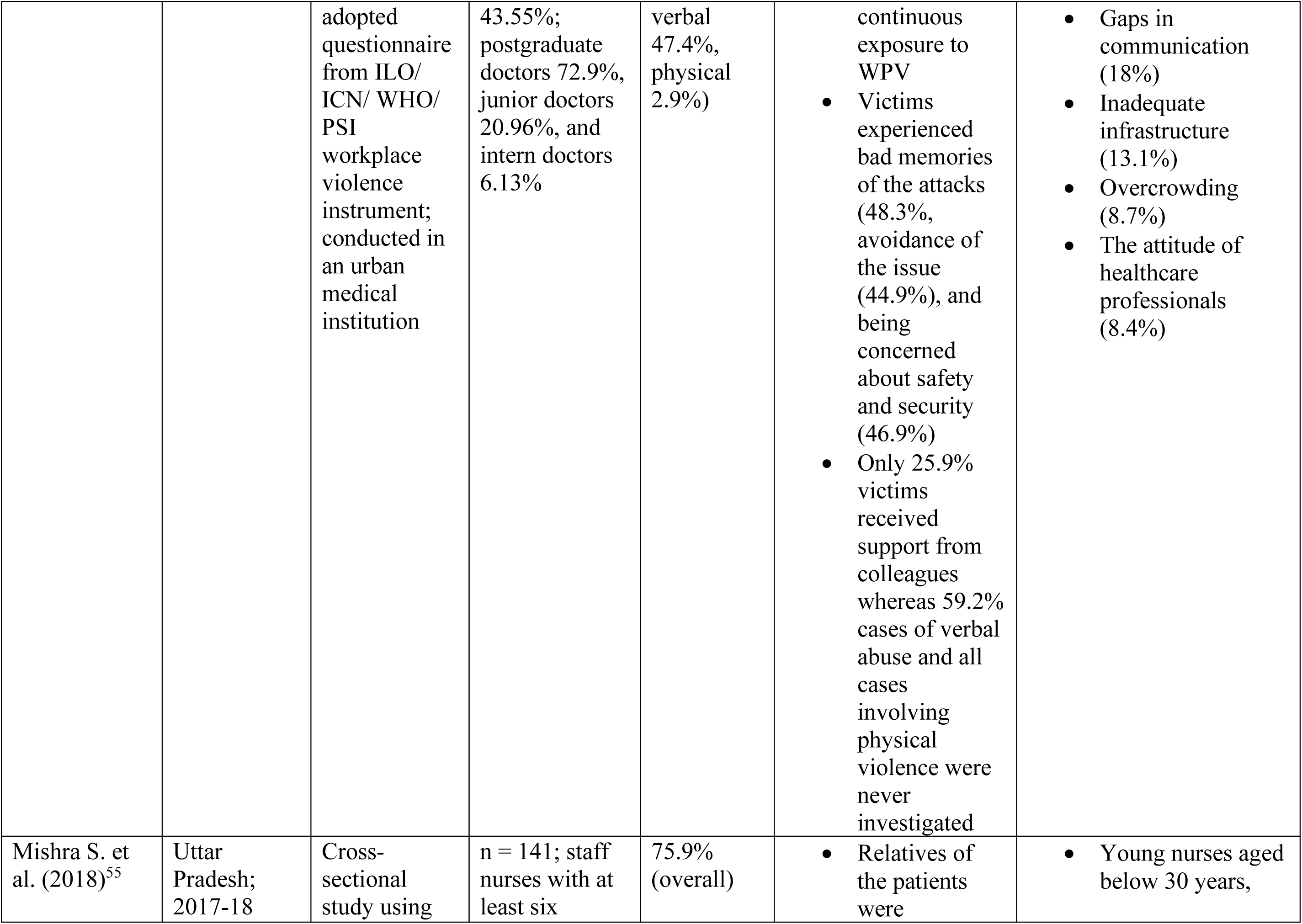
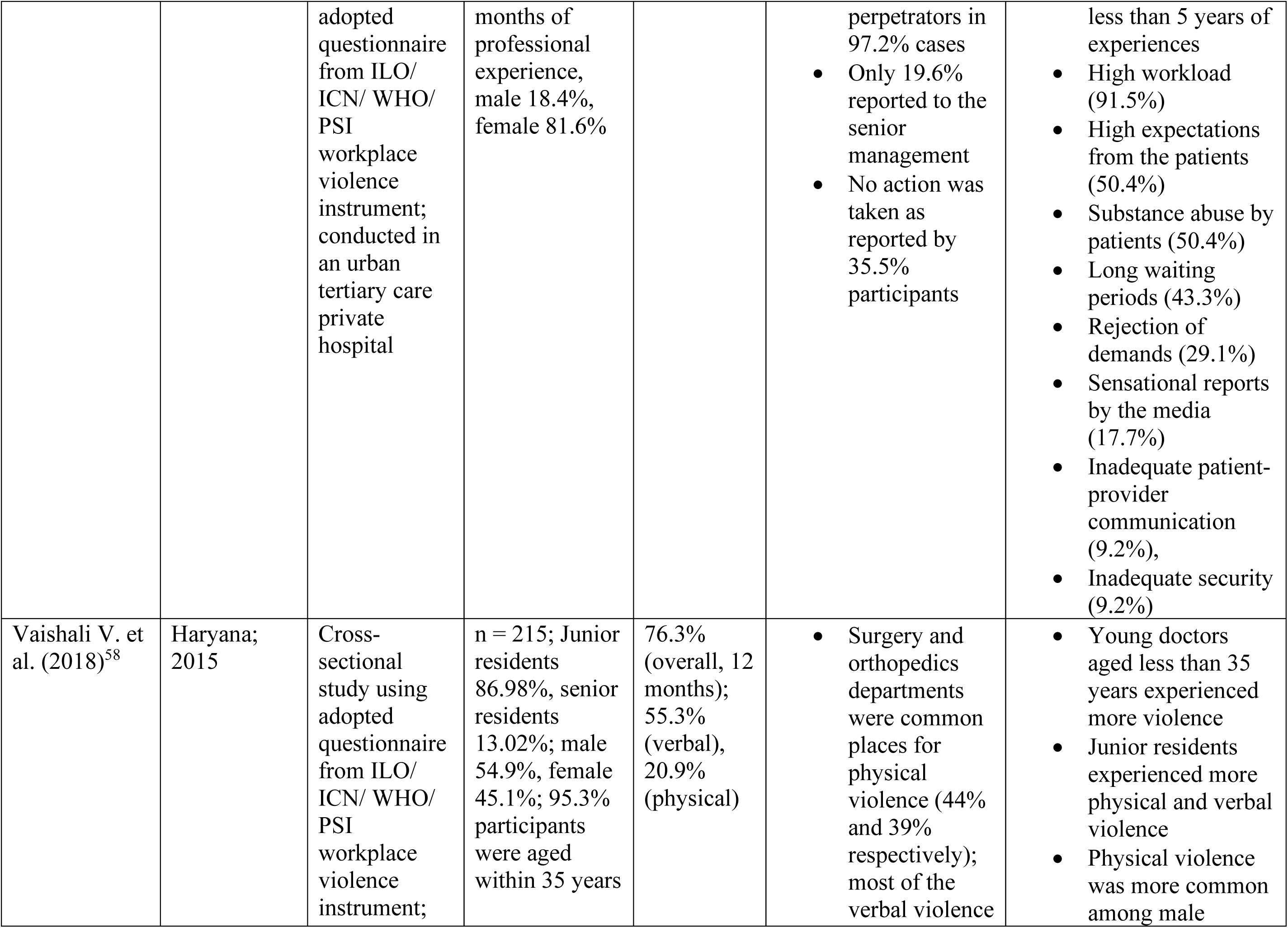
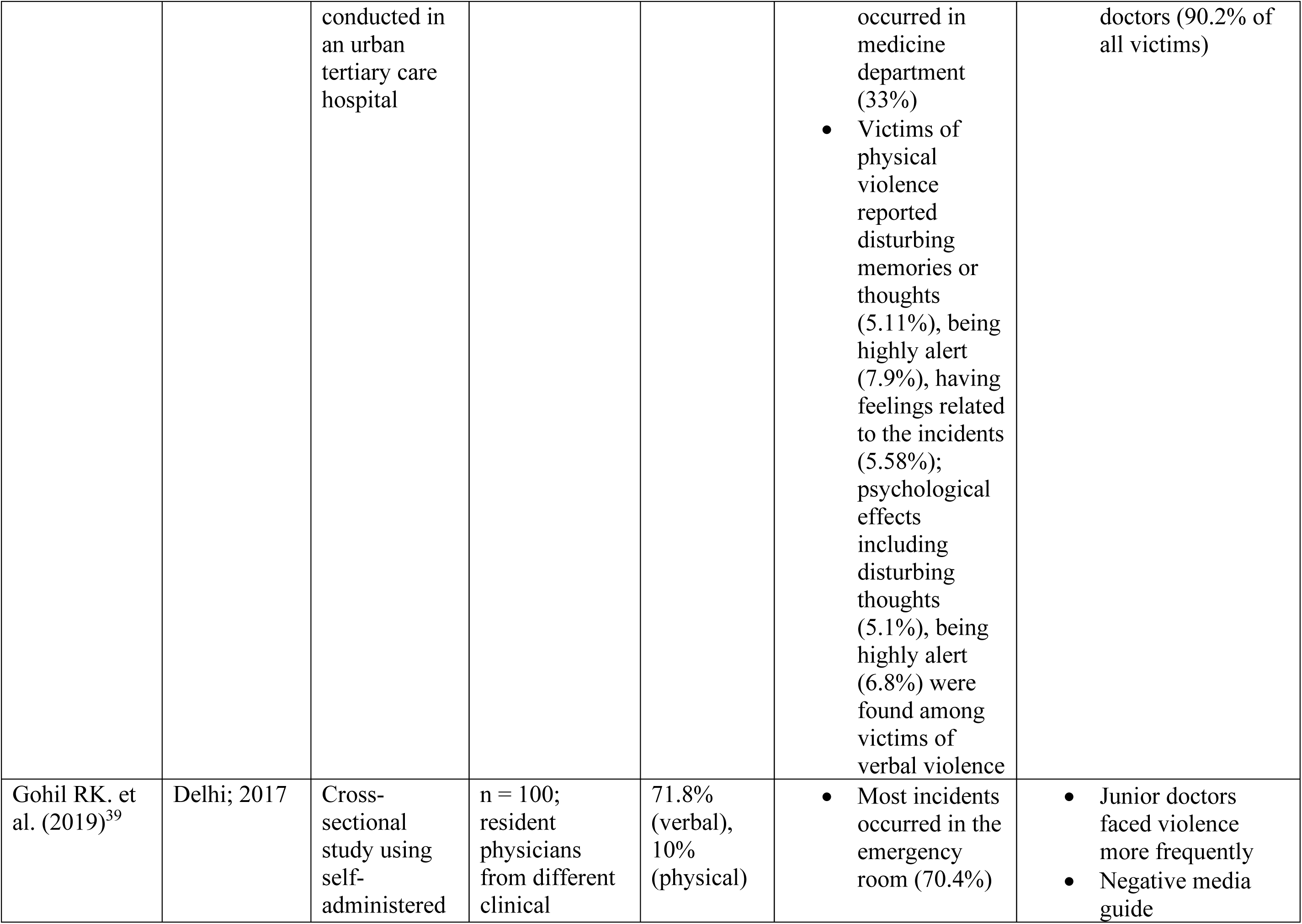
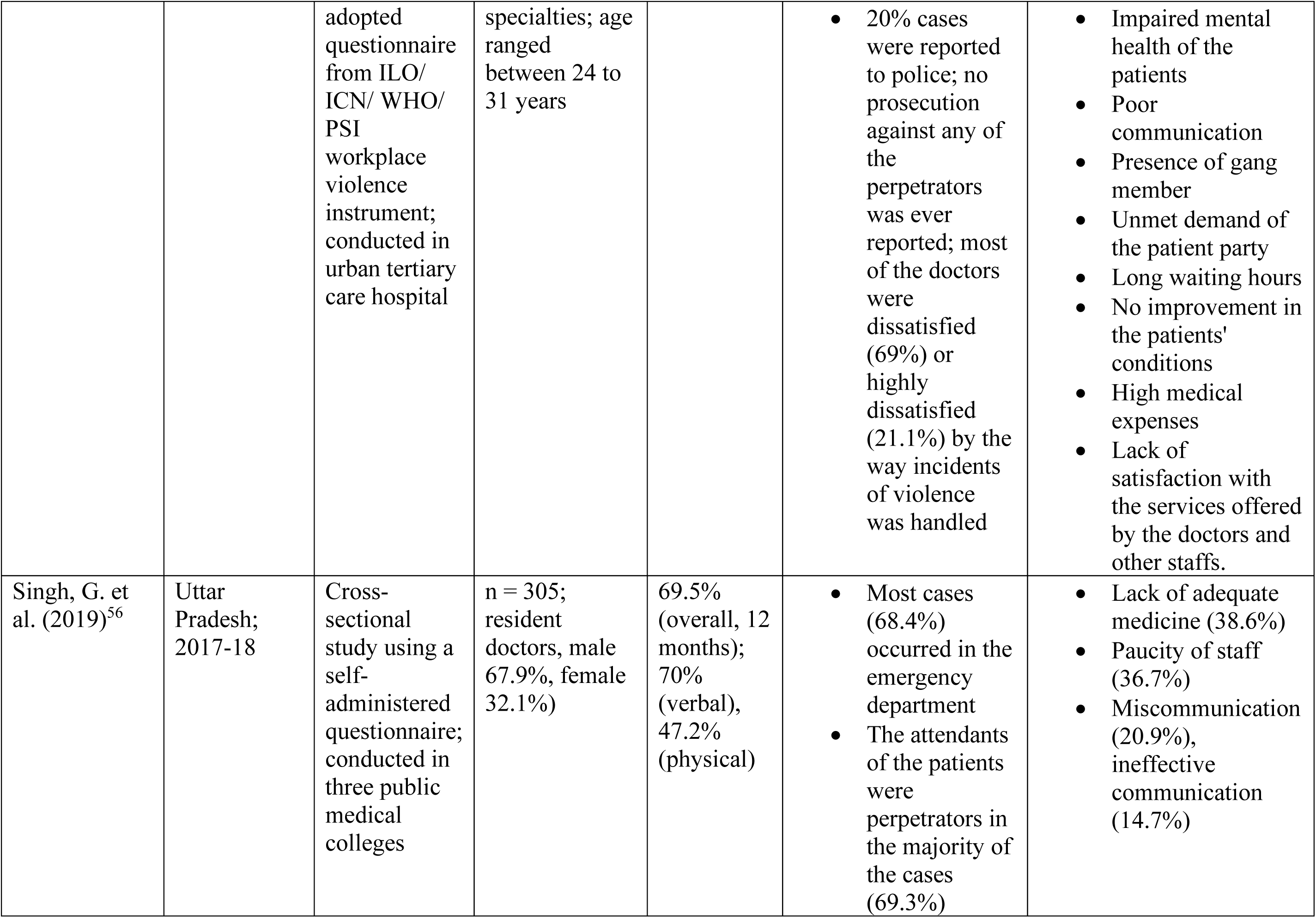
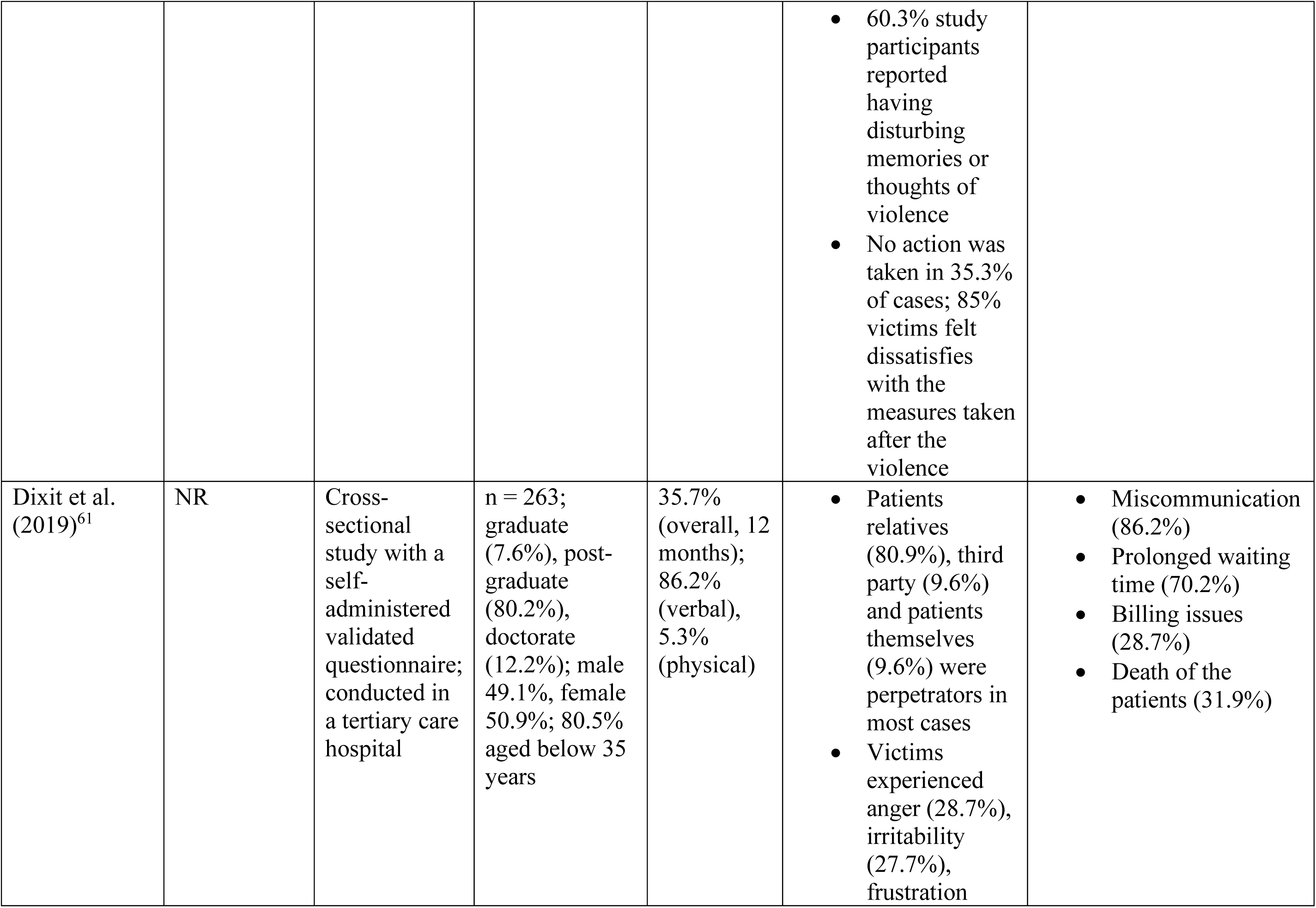
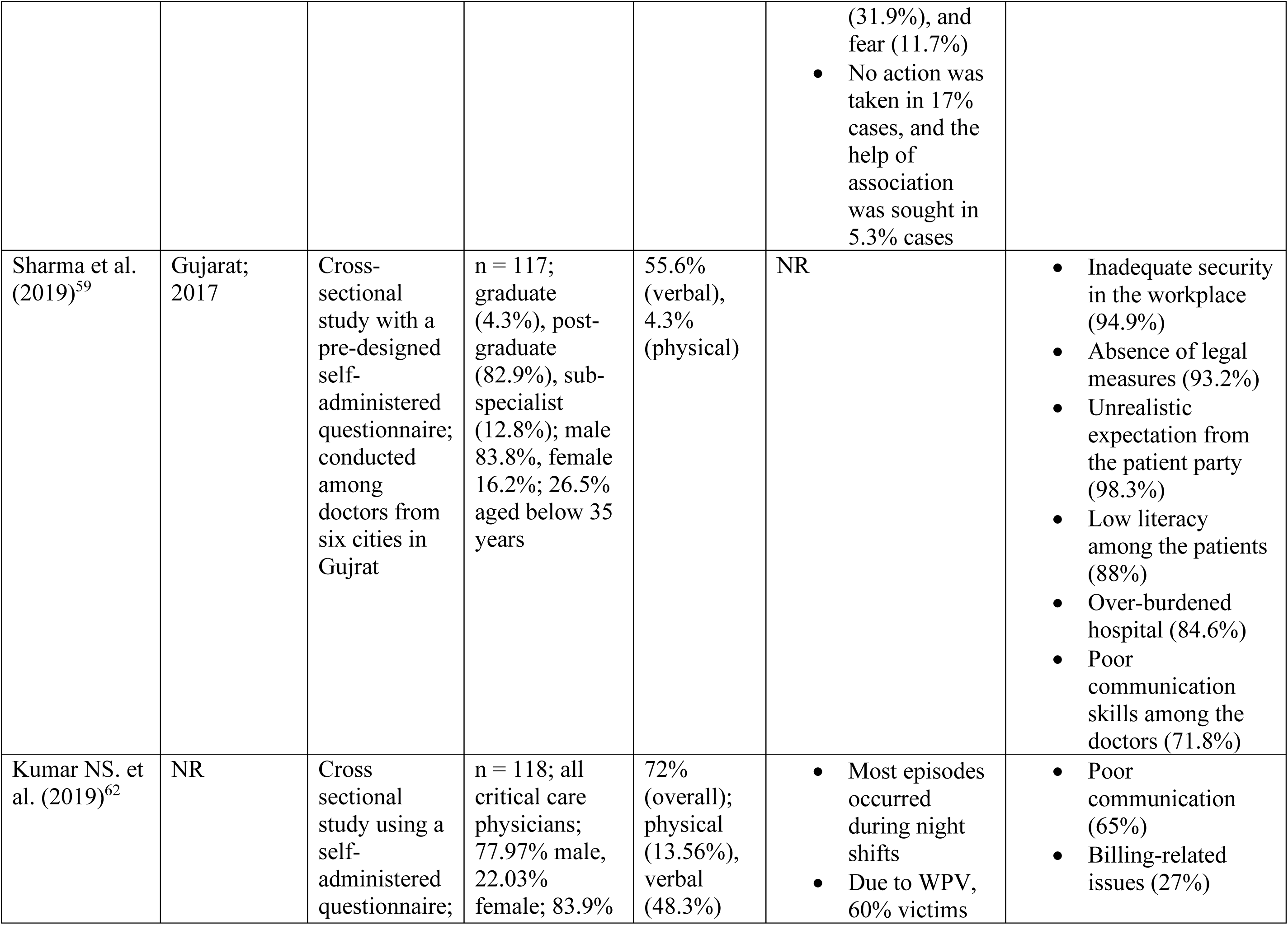
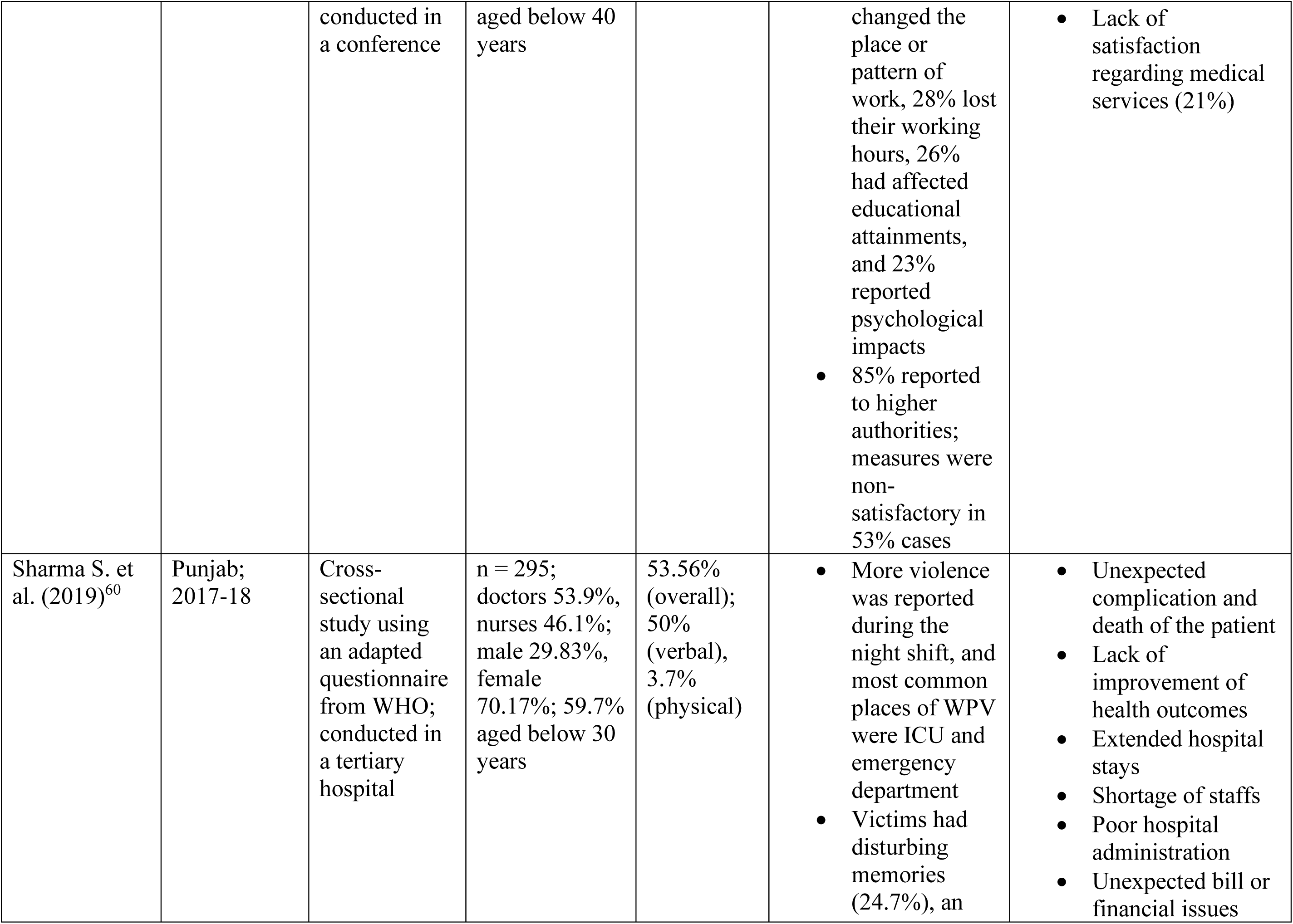
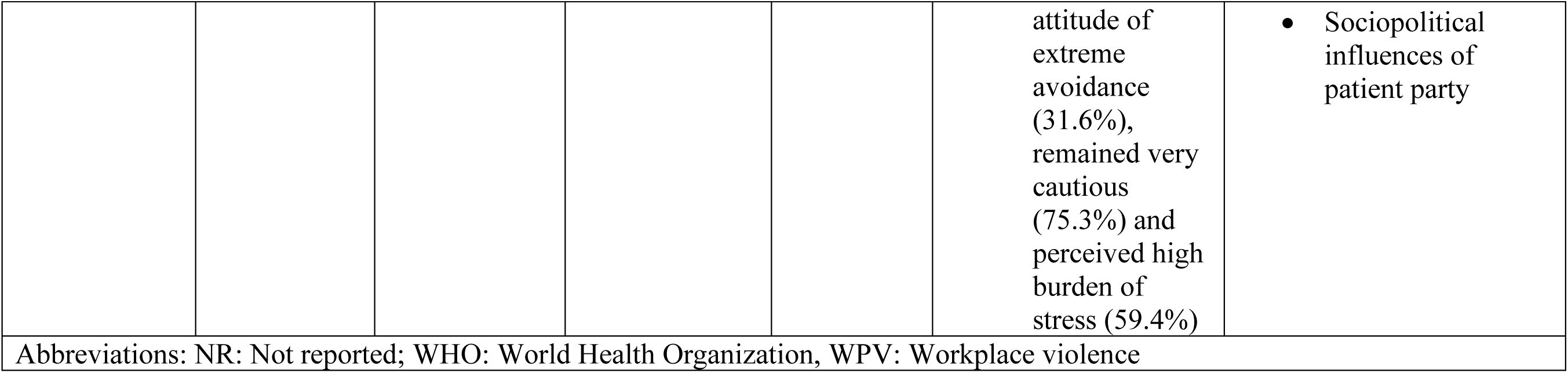
Summary of the Studies Included in this Review.

**Figure 1:**
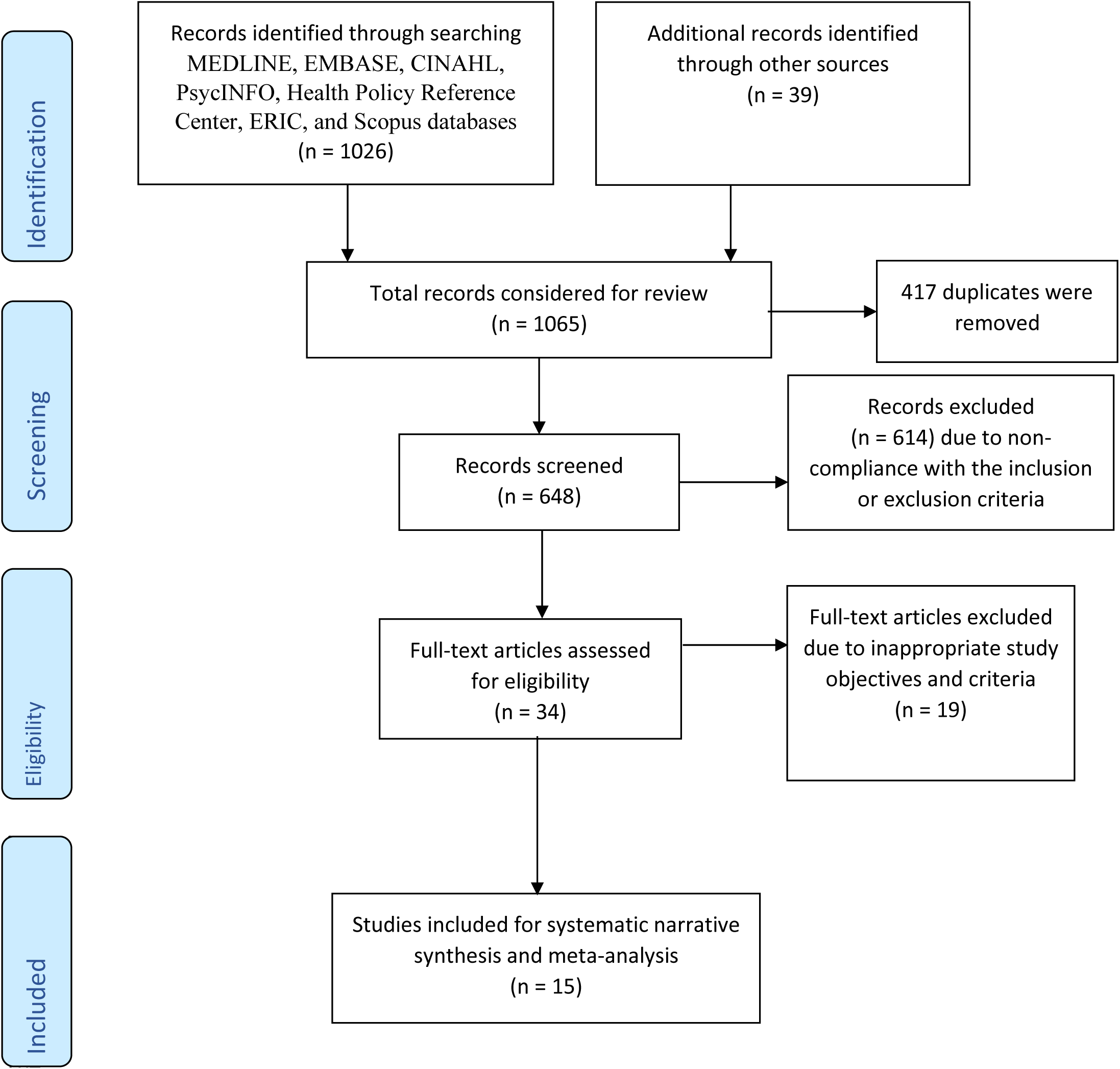
Flow Diagram of the Systematic Review Process.

### 3.1 Characteristics of the Recruited Studies

Among the included articles, earlier studies were conducted in Tamil Nadu (n = 1)^51^ and Karnataka (n = 1)^52^ in 2005 and 2006 respectively. Most of the studies (n = 13 out of 15) were conducted after 2010 in different states of India including Delhi (n = 3),^39–41^ Manipur (n = 2),^53,54^ Uttar Pradesh (n = 2),^55,56^ Maharashtra (n = 1),^57^ Haryana (n = 1),^58^ Gujarat (n = 1),^59^ and Punjab (n = 1).^60^ The quality evaluation using the NHLBI tool found no studies with poor quality, three studies with fair quality,^39,51,57^ and remaining studies (n = 12) with good quality.^40,41,61,62,52–56,58–60^

Most of the studies (n = 14) recruited study participants from medical institutions in urban areas whereas only one study by Pund et al. recruited from multiple locations in a district where 65.85% participants were working in rural areas.^57^ All these studies were cross-sectional in nature and conducted using self-administered pre-tested questionnaires with different demographic and violence-related variables. While most of the studies (n = 14) reported the magnitude of violence in percentage, one study by Balamurugan reported the findings using violence score among the victims.^52^ Five studies particularly used adopted questionnaire from joint workforce violence instrument developed by International Labor Organization (ILO), International Council of Nurses (ICN), World Health Organization (WHO), and Public Services International (PSI).^39,54,55,58,60^

Fifteen studies recruited in this study had a varying number of study participants ranging from 82 to 310, totaling 2849 participants. Most of the studies (n = 13) had a majority of male participants except three studies,^52,55,60^ which comprised nursing professionals. All the remaining studies (n = 12) had participants including resident physicians, intern physicians, and other cadres of healthcare professionals.

### 3.2 Prevalence of WPV Against Healthcare Professionals in India

In the meta-analysis, the pooled prevalence of WPV was found as 63% (95% CI, 55%-72%) among 2849 participants from the 15 recruited studies in this review (Figure 2). There was highly statistically significant heterogeneity (I^2^ = 95.79%, *p* = 0.00) for which the random-effects model was used in this analysis.

**Figure 2:**
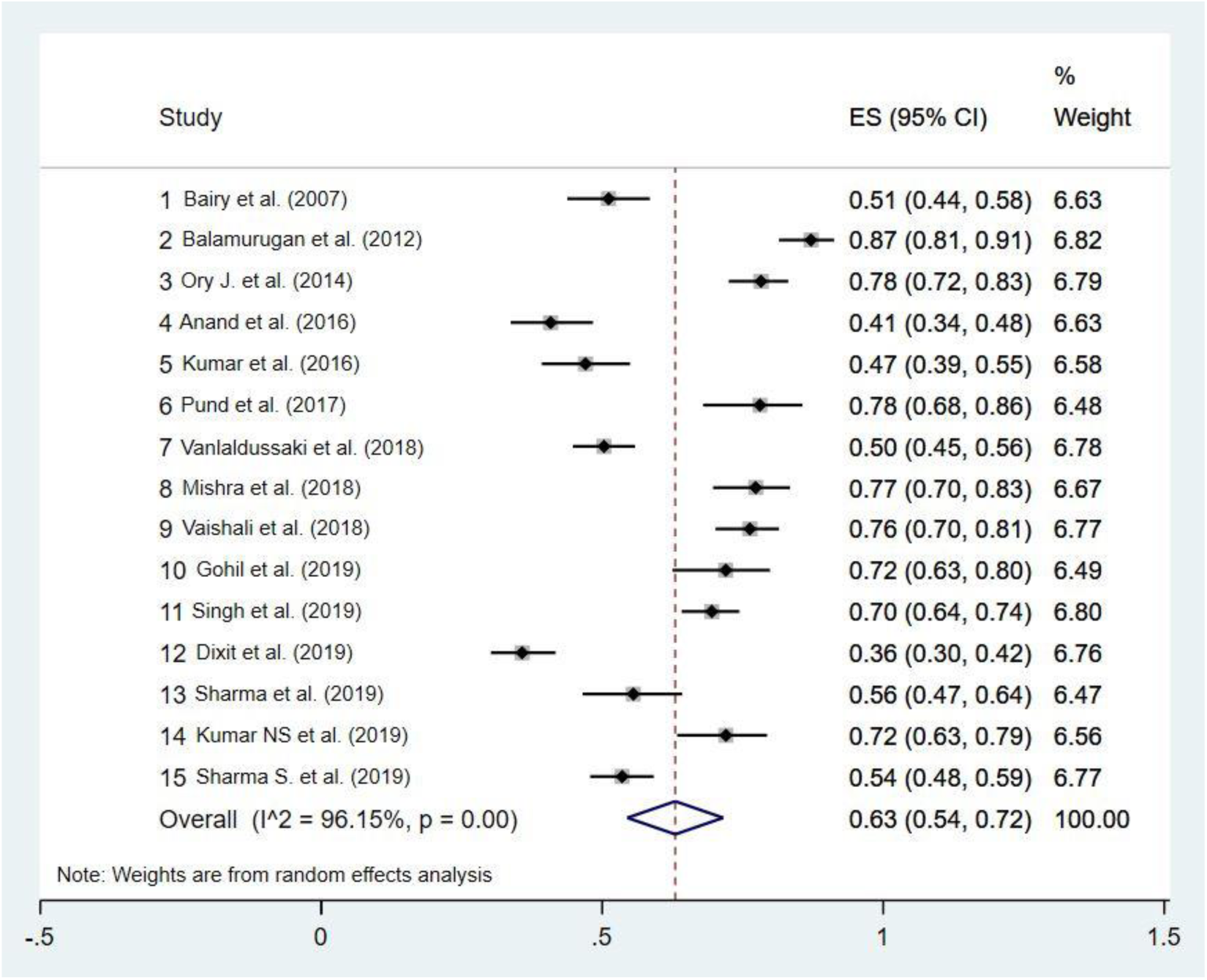
Pooled Prevalence of WPV Against Healthcare Professionals in India.

In the subgroup analysis, pooled prevalence for variables including gender (male or female), sample size (less or more than 200), publication year (studies published before or after 2017), timeframe of prevalence reporting (12-months or lifetime), and types of violence (verbal or physical) were estimated (Table 3). Nine studies reported gender-specific cases of the prevalence of WPV for male and female, which revealed a pooled estimate of 64% (95% CI, 51%-78%) and 54% (95% CI, 47%-62%) among male and female participants respectively.

**Table 3:** Subgroup Analysis of the Prevalence of WPV.

| **Variables** | **Number of studies** | **Number of participants** | **Prevalence (95% CI)** | **I<sup>2</sup> value** | **p value** |
| --- | --- | --- | --- | --- | --- |
| *All* | 15 | 2849 | 63% (54-72) | 96.15% | 0.00 |
| ***Gender*** |  |  |  |  |  |
| Male | 9 | 921 | 64% (51-78) | 95.90% | 0.00 |
| Female | 9 | 846 | 54% (47-62) | 78.60% | 0.00 |
| ***Sample size*** |  |  |  |  |  |
| Less than 200 | 9 | 1231 | 65% (53-76) | 95.50% | 0.00 |
| More than 200 | 6 | 1618 | 61% (47-74) | 97.10% | 0.00 |
| ***Publication year*** |  |  |  |  |  |
| Published till 2017 | 6 | 985 | 64% (48-80) | 97.10% | 0.00 |
| Published after 2017 | 9 | 1864 | 62% (53-72) | 95.20% | 0.00 |
| ***Timeframe of prevalence estimation*** |  |  |  |  |  |
| 12-months | 7 | 1290 | 55% (43-67) | 95.50% | 0.00 |
| Lifetime | 9 | 1641 | 69% (61-78) | 93.80% | 0.00 |
| ***Type of violence*** |  |  |  |  |  |
| Verbal violence | 14 | 2780 | 52% (45-60) | 93.90% | 0.00 |
| Physical violence | 13 | 2496 | 8% (5-11) | 93.70% | 0.00 |
| Abbreviations: WPV: Workplace violence; CI: Confidence interval |  |  |  |  |  |

Nine studies with sample size less than 200 had a pooled prevalence of 65% (95% CI, 53%-76%) whereas six studies with more than 200 samples had a pooled prevalence of 62% (95% CI, 47%-74%). Moreover, six studies were published until 2017, which had a pooled prevalence of 64% (95% CI, 48%-80%). Nine studies published after 2017 had a pooled prevalence of 62% (95% CI, 53%-72%). Studies reported the prevalence of WPV in different timeframes. For seven studies reporting a 12-month prevalence of WPV, the pooled prevalence was 55% (95% CI, 43%-67%) whereas nine studies reported a lifetime prevalence of WPV with a pooled prevalence of 69% (95% CI, 61%-78%). Furthermore, the pooled prevalence of verbal and physical violence was 52% (95% CI, 45%-60%) and 8% (95% CI, 5%-11%), respectively.

### 3.3 Characteristics of the WPV

#### 3.3.1 Location, Time, Frequency, and Magnitude of WPV

Six studies reported emergency department as the most commonplace of WPV.^39,40,53,56,57,60^ For example, Pund et al. reported that the doctors working in emergency department were more likely to experience WPV (odds ratio [OR] 4.77, 95% CI 1.45-15.63).^57^ Other places of occurring WPV included department of medicine,^41,58^ surgery,^41,58^ gynecology and obstetrics,^41^ and intensive care unit.^60^ Two studies reported evening and night shifts as the common time when most incidents of WPV were observed.^59,62^ Moreover, studies have also reported varying magnitudes of violence. For example, Ori et al. reported 38.89% participants experienced WPV for three or more times.^53^ Another study by Vanlalduhsaki et al. reported 8.8% participants had persistent exposure to WPV.^54^ Furthermore, the magnitude of violence was diverse in a few studies involving gender-based violence alongside physical and verbal violence. For example, Ori et al.^53^ reported one case of sexual violence and Gohil et al.^39^ reported sexually offensive verbal violence against the study participants, which involved both physical and verbal abuses. Also, participants experiencing the same type of violence had varying levels of exposure to the WPV. For example, Balamurugan et al. reported the level of violence as mild and moderate among 57% and 30.2% participants, respectively.^52^

#### 3.3.2 Impacts of WPV Among the Victims

Incidents of WPV had multiple consequences among the victims. Six studies reported varying levels of psychological impacts among the victims of WPV.^40,54,56,58,60,62^ For example, Anand et al. reported fear (59%), sadness (44.3%), headache (31.1%) and other psychological problems after WPV.^40^ Disturbing memories of WPV affected the victims as reported in four studies.^54,56,58,60^ Three studies reported a more cautious attitude in the workplace and avoidance of WPV-related discussions among the victims.^54,58,60^ Victims also reported WPV affected their self-esteem and professional confidence.^51^ Moreover, Pund et al. reported extorsion and intentional destruction of property among the cases of WPV.^57^ Studies conducted by Vanlalduhsaki et al. and Pund et al. reported a diminished sense of safety in the workplace as found among the victims of WPV.^54,57^ Furthermore, a study by Kumar NS et al. reported an altered working pattern and lost working hours among the victims of WPV.^62^

#### 3.3.3 Perpetrators of WPV and Measures Following WPV

Six studies reported the perpetrators of WPV.^40,51,53,55,56,61^ In most studies (n = 5),^40,53,55,56,61^ the relatives or attendants of the patients were major perpetrators of WPV whereas two studies reported the involvement of hospital staffs in abusing their colleagues.^40,51^ In most cases, the incidents of WPV was not reported to higher authorities. Low reporting was documented in seven studies which mentioned about a reporting system.^39–41,51,55,61,62^ Moreover, the actions taken by the respective authorities were limited as revealed in studies conducted by Pund et al.,^57^ Vanlalduhsaki et al.,^54^ and Kumar M. et al.^41^ In addition, Kumar NS et al.,^62^ Singh et al.,^56^ and Gohil et al.^39^ reported the victims were not satisfied with the adopted measures following WPV. In studies conducted by Dixit et al.,^61^ Gohil et al.,^39^ and Mishra et al.,^55^ the participants reported that no action was taken following the incidents of WPV.

### 3.4 Factors Associated with WPV

Several factors associated with WPV were reported across studies, among which problems related to patient-provider communication were reported in most of the studies (n = 10).^39,40,53– 57,59,61,62^ For example, Ori J. et al. reported difficulties in communicating between doctors and patients contributed to WPV.^53^ Another study by Anand et al. highlighted poor communication skills among 81.1% of the participating healthcare providers, which was identified as a critical factor associated with WPV.^40^ Another key factor associated with WPV was a lack of satisfaction among the patients (n = 7).^39,40,55,59–62^ For example, a study by Gohil et al. reported that the patients were not satisfied with the services offered to them, which aggravated incidents of WPV.^39^ Moreover, many studies (n = 7) reported fewer years of professional experience among the providers was associated with WPV.^39,41,51,52,55,57,58^ For example, Pund et al. reported that young doctors with less experience in workplaces were more likely to experience WPV compared to their senior colleagues (OR 3.67, 95% CI 1.28-10.47).^57^ Moreover, inadequate resources and poor infrastructure affected the overall working environment and contributed to WPV, as reported in six studies.^40,53,54,56,57,60^ For example, Anand et al. reported a shortage of resources as a major reason (73.4%) of WPV.^40^ Another factor contributing to WPV was long waiting hours in the hospitals as reported in five studies.^39–41,55,61^ In a study by Kumar M. et al., 73.5% of the participating healthcare providers reported long waiting hours as a challenge which might have influenced the patients and their relatives in the cases of WPV.^41^

Further, five studies have reported excessive workload and overcrowding of the hospitals affecting the health service delivery and increasing the incidents of WPV in the respective hospitals.^40,53–55,59^ Anand et al. reported overcrowding was perceived as a critical factor related to WPV by 77.5% participants.^40^ Another factor associated with WPV was issues related to billing and medical expenses, which was reported in four studies.^39,60–62^ For example, Dixit et al. reported billing was associated with WPV as identified by 28.7% participants,^61^ which were similar to the findings of Kumar NS. et al. where 27% participants reported medical expenses were associated with WPV.^62^ Moreover, four studies reported a lack of protective measures to make the workplace secure against WPV.^53,55,59,60^ A study by Sharma et al. reported inadequate security (94.9%) and the absence of legal measures (93.2%) to protect the health workforce from WPV and its consequences.^59^ Most of the studies reported aggregated data for all genders; however, Ori J. et al. and Vaishali et al. reported WPV was higher among male participants.^53,58^ In contrast, Balamurugan reported higher incidents of WPV among the female participants.^52^ Other factors associated with WPV included behavioral issues of the patients and the healthcare professionals (n = 3),^39,54,55^ lack of literacy among the patients and their relatives (n = 2),^53,59^ death of the patient (n = 2),^60,61^ socio-political influences of the patients (n = 2),^39,60^ and negative reports in mass media (n = 2).^39,55^

## 4. Discussion

This review evaluated empirical studies and reported the current evidence on WPV. In the context of India, a wide range of health systems challenges deteriorate the desired roles, activities, and outcomes in healthcare organizations resulting in unexpected incidents like WPV. In addition, incidents of WPV can critically affect the relationships between patients and their healthcare providers, the wellbeing of the patients as well as the health workforce, delivery of health services, trust on the health system, and overall quality of care. Therefore, WPV can be examined as a vicious process within the resource-constrained health system in India. It is essential to acknowledge how individual, interpersonal, organizational, and systems-level factors interact with each other and contribute to WPV. This review identified the characteristics and associated factors of WPV, which show the complex structural and functional challenges within the existing health services organizations. These findings offer several insights which are critical for future research, policymaking, and practices. To our knowledge, this is the first systematic review and meta-analysis presenting the prevalence, characteristics, and associated factors of WPV against healthcare professionals in India.

In this review, most of the studies reported WPV in tertiary hospitals, which are mostly located in urban areas. Therefore, studies in this review do not inform the severity of WPV in rural areas of India. Nearly 68.84% of Indian population live in rural areas which have less-developed infrastructures, a more severe paucity of human resources, low literacy among the general population, high poverty and inability to pay for health services, and many other barriers to deliver health services.^63^ In addition, the rural health system largely depends on community-level health workforce, including Accredited Social Health Activists and Anganwadi workers who have lesser skills and resources compared to healthcare professionals in urban centers.^64,65^ These challenges increase the likelihood of WPV in rural areas. Moreover, the magnitude and attributable factors of WPV can be different in private healthcare organizations. These areas are not adequately explored in the existing studies. Future research should address these knowledge gaps and explore the severity and underlying factors associated with WPV in different geographic and organizational contexts within India.

Another concept in this discourse is the patient-provider communication (PPC), which plays a critical role in patient-centered decision-making, accessing and utilizing healthcare services, and achieving desirable health outcomes.^66^ Ineffective PPC can serve as a major barrier in healthcare organizations leading to inadvertent conditions like WPV.^67,68^ Most studies in this review identified inadequate and ineffective PPC as a major challenge necessitating further assessment and actions. There is a lack of empirical studies discussing the determinants of PPC in the socio-cultural context of India.^69^ Moreover, the existing health education programs offer little emphasis on socio-behavioral aspects of formal caregiving in healthcare professions.^70^ These challenges suggest that there is a critical lack of knowledge and preparedness among the providers on establishing and managing meaningful PPC in their workplaces. Educational and psychosocial interventions for healthcare providers can help in overcoming this crisis and enable them to better communicate with their patient and manage health problems with empathy.^71,72^

Moreover, several challenges including low literacy, poverty, behavioral issues, and dissatisfaction are reported among patients attending healthcare organizations in India. Their problems require careful attention from both the healthcare and social perspectives. In low- and middle-income countries, socio-behavioral challenges among the patients often lead to suboptimal participation in health services.^73^ Increasing awareness and engaging the patients in health services would necessitate behavior change communication in both healthcare and community levels.^74^ However, addressing social inequalities would need a broader approach where the healthcare providers can collaborate with social welfare agencies to alleviate social and structural barriers, resolve the perceived challenges specific for contexts and people, and address socio-behavioral risk factors of WPV in a given population.^72,74^

It is essential to recognize that a shortage of resources in a healthcare organization was revealed as a major challenge related to WPV. In India, most of the public hospitals suffer from a severe scarcity of healthcare providers.^75^ Moreover, the inadequate supply of medication and a lack of standard operating procedures for clinical and non-clinical activities in the hospitals result in suboptimal health services and outcry among the patients.^76^ Policymakers and healthcare leaders should acknowledge this gap and establish institutional approaches to assess the logistic problems and meet the institution-specific demands in a timely manner. Furthermore, scarcity of human resources should be addressed through better human resource strategies including recruitment of adequate human resources, educating and preparing them for optimal service delivery, deploying them ensuring equitable distribution, and creating an enabling environment where they can serve with satisfaction and excellence.^77^ In addition, recruiting interpreters and public relations personnel can facilitate health communications and service delivery, which may alleviate the workload of the healthcare professionals and improve holistic care in healthcare organizations.^53^

Furthermore, poor and unsafe infrastructure critically affects the overall delivery and utilization of health services.^78^ In resource-constrained contexts, poorly designed healthcare facilities can foster discomfort and impatience among patients and healthcare professionals.^78,79^ It is essential to consider such structural issues while designing and improving healthcare infrastructures. In addition, overcrowding is common among public hospitals in India, where each tertiary hospital is likely to have manifolds of patients compared to the institutional capacities.^53,54^ This leads to overutilization of the resources with minimal repairing or restoration of the infrastructures and amenities. This is more of a system’s challenge where the health systems should be strengthened by improving the structural and functional capacities of the healthcare infrastructures to meet the population health needs.^78^

In such a scenario, healthcare security management and surveillance systems supported by advanced information technologies can help not only in oversight of the security of the organizations but in making decisions to prevent potential conflicts.^80,81^ Such systems may include real-time video surveillance of the hospital areas, monitoring the movement of individuals to predict and prevent unlawful activities, maintaining digital records of the patients, visitors, employees, and other individuals entering in the hospital premises.^81,82^ In addition, such surveillance systems can be integrated within Health Management Information System (HMIS) at the local, states, and national levels; which may allow stakeholders at different levels and agencies to take preventive and corrective measures to make safer healthcare organizations.

This review also found that the emergency department was a commonplace of WPV in India, which requires an in-depth analysis of the underlying problems in the emergency management and referral systems in Indian healthcare. In most areas of India, the emergency response system is inadequate to deal with mass casualties from incidents like road traffic accidents, natural or man-made disasters, and other emergency conditions requiring coordinated care.^83^ Moreover, the primary care centers are generally ill-equipped to manage emergency cases, which are generally referred to urban multi-specialty hospitals.^84^ This continuous inflow of patients often leads to overcrowding near the emergency department of urban hospitals, which has pre-existing limitations and challenges.^83^ Understanding this cycle of emergency management can better inform how potential strategies can be developed and implemented to strengthen emergency management and referral services. Such strategies may include hotlines for rapid response in emergency cases, ambulance services for mobilizing the patients, institutionalizing protocols for trauma management guiding specific types of patients to pre-specified centers without delay, and establishing a centralized information system for coordinating the patients and health services organizations for better emergency and referral care.^83,85^

Many studies in this review reported critical gaps in existing systems of reporting WPV and insufficient measures taken by the hospital authorities, which may affect the occupational engagement and personal wellbeing of the victims of WPV. Also, the physical and mental health of the victims of WPV should be taken care of alongside the administrative measures.^55,56^ Without addressing these challenges, psychosocial and structural violence against the victims may appear to be more severe than WPV itself. Health services organizations should acknowledge their accountability to ensure the safety at the workforce and institutionalize pro-active systems of reporting and investigating the incidents of WPV.

Furthermore, addressing WPV would require a legislative perspective to analyze the same in the context of existing policies and laws within healthcare organizations, respective states, and the nation. In India, 19 states have laws for protection of medical professionals and healthcare establishments, which are essential to enforce in practice to prevent and address WPV.^86^ Legal and administrative challenges to operationalize existing legislative measures should be explored and future policies should be developed considering the high societal burden of WPV against health workforce in India.

Lastly, a centralized health system may affect participatory planning and implementation of programs and policies to address WPV at the local level. Moreover, factors associated with WPV can be unique from one place to another. Therefore, engaging stakeholders at the local and regional levels and considering their collective perspectives can facilitate better policymaking and preventing WPV. Such coordination among stakeholders in different levels may protect the healthcare providers and enable them to work safely in central and remote healthcare organizations.

This systematic review has several limitations which are essential to acknowledge. One such limitation is the biases within the existing body of literature. We included studies from specific databases and additional sources; however, it is possible that there are studies which might have met our criteria but did not appear in our searching process. Such studies, if there are any, could have affected the findings of the review. Moreover, publication bias may exist within the studies. As observed in prevalence meta-analyses, there is a lack of consensus on the methods of evaluating publication bias for prevalence studies.^87^ Conventional funnel plots may provide spurious asymmetry despite having no publication bias.^88^ Acknowledging this limitation of application and interpretation, we did not analyze publication bias in this meta-analysis. Furthermore, various factors associated with WPV were reported across studies which had high heterogeneity in methods and itemized responses, which did not allow to meta-analyze specific risk factors; which is another limitation of this review. Future research should acknowledge the above-mentioned limitations and inform better evidence with higher precision.

## 5. Conclusion

This systematic review and meta-analysis evaluated the current evidence and found a high prevalence of WPV against healthcare professionals in India. Moreover, the characteristics and associated factors of WPV in the empirical studies inform the magnitude and underlying reasons of WPV in the complex healthcare and socio-cultural context of India. Limitations in the existing literature necessitate further research with rigorous design examining different aspects of WPV among the healthcare professionals. However, the current evidence reported in this systematic review informs a need to acknowledge the high burden of WPV experienced by the healthcare professionals, address the factors associated with WPV, create an safe and enabling environment in healthcare organizations, and protect the healthcare professionals so that they can protect the health and wellbeing of their patients without the fear of WPV.

## Data Availability

All data related to this systematic review and meta-analysis is available upon request.

## Acknowledgements

None.

## Notes

### Competing Interest Statement

The authors have declared no competing interest.

### Funding Statement

No funding was received at any stage of conducting this review or preparing this manuscript.

